# Concerns, quality of life, access to care and productivity of the general population during the first 8 weeks of the coronavirus lockdown in Belgium and the Netherlands

**DOI:** 10.1101/2020.07.24.20161554

**Authors:** Hanne van Ballegooijen, Lucas Goossens, Ralph H. Bruin, Renée Michels, Marieke Krol

## Abstract

**Background:** The COVID-19 pandemic has a disruptive impact on our society. We therefore conducted a population survey to describe: 1) stress, concerns and quality of life 2) access to healthcare and cancelled/delayed healthcare and 3) productivity during the first 8 weeks of the coronavirus lockdown in the general population.

**Methods:** An online survey was conducted in a representative sample after 8 weeks of the coronavirus lockdown in Belgium and the Netherlands. The survey included questions about stress, concerns, quality of life delayed/cancelled medical care and productivity loss using validated questionnaires.

**Results:** In total, 2099 Belgian and 2058 Dutch respondents completed the survey with a mean age of 46.4 and 42.0 years, respectively. Half of the respondents were female in both countries. A small proportion tested positive for COVID-19, 1.4% vs 4.7%, respectively. The majority of respondents with a medical condition was worried about their current health state due to the pandemic (53%) vs (63%), respectively. Respondents experienced postponed/cancelled care (26%) and were concerned about the availability of medication (32%) for both countries. Productivity losses due to the COVID-19 restrictions were calculated in absenteeism (36%) and presenteeism (30%) for Belgium, and (19%) and (35%) for the Netherlands. Most concerns and productivity losses were reported by respondents with children <12 years, respondents aged 18-35 and respondents with an (expected) COVID-19 infection.

**Conclusions:** This study describes stress, quality of life, medical resource loss and productivity losses in Belgium and the Netherlands after 8 weeks of coronavirus lockdown. The results underline the burden on society.

## Introduction

The COVID-19 pandemic has had an enormous disruptive impact on societies throughout the world (1, 2), not only because of its direct impact on patients with COVID-19, but also due to fear and stress (3). These include fears for unemployment due to COVID-19-related restrictions, financial worries and concerns about one’s own health. These stressors and worries may be reflected in lower quality of life.

Next to the impact of the COVID-19 related restrictions on quality of life, concerns and stress, the COVID-19 pandemic also caused a substantial strain on the healthcare system (4). To cope with the COVID-19 infected patients, healthcare facilities had to cancel regular care. In addition, patients may have been anxious to visit to their physician or general practitioner because of fear of infection or to avoid further burdening the healthcare system. This could lead to large, secondary healthcare problems such as lower number of diagnoses of critical diseases and exacerbation of existing health conditions without medical intervention.

Additionally, the COVID-19 pandemic has had a wider impact on social and economic factors (5, 6). Governments have imposed measures aimed at reducing the spread of the virus. These changes imposed on society impacted individuals and organizations. As a result, many individuals and organizations were forced to change their normal routines. Examples are closing of schools and day-care centers, restaurants and sport centers, and asking people to work from home. Many individuals who needed to work from home were forced to combine work with childcare. These measures may have impacted individuals’ productivity, as people may not have been able to work at their normal capacity. Not being able to work at normal capacity while at work (either at home or in the office) is referred to as presenteeism. Others may not have been able to work at all, for instance those working in restaurants or sport centers. Not being able to do one’s work at all is referred to as absenteeism.

Although the combination of presenteeism and absenteeism may have had a widespread impact on society, there is little scientific information on these effects in the general population. Therefore, the aim of this study was to observe and record the developments during the coronavirus lockdown in Belgium and the Netherlands. The governments of Belgium and the Netherlands imposed a lockdown in early March 2020 to prevent further spread of the coronavirus. In Belgium, the recommended distance was 1.5 meters. In addition, it was forbidden to go outside if not strictly necessary and schools and universities were closed (7). In the Netherlands, individuals could leave their home, but were asked to stay at home as much as possible. They also needed to keep 1.5 meters distance. Schools, day-care facilities, restaurants and sport centers were closed in both countries.

In May 2020, after 8 weeks of the coronavirus lockdown, we conducted a population survey to gain insight into the following aspects 1) stress, concerns and quality of life 2) access to healthcare and cancelled/delayed healthcare and 3) productivity in Belgium and the Netherlands.

## Methods

### Study design and participants

A cross-sectional, web-based survey was conducted in Belgium and the Netherlands in the beginning of May 2020. At that time, both Belgium and the Netherlands had experienced 8 weeks of COVID-19-related restrictions. Data were collected over a time span of 1 week in a large online panel by sampling agency Dynata. The aim was to include a sample of 2000 respondents from both countries of 18 years and older with a representative spread for age, sex, education and region. The Belgian sample consisted of equal proportions of Dutch and French speakers.

The data collection was in accordance with local and European privacy legislation. Respondents’ identity remained unknown to researchers and all respondents provided informed consent regarding their participation in the survey and for scientific publication.

### Questionnaire

The questionnaire was developed in Dutch and translated into French. The questionnaire was pilot tested among 5 participants and after a soft launch involving 100 respondents’, small adjustments were implemented to improve the answer categories. The questionnaire started with demographic questions, which included age, sex, marital status, number and age of children, and education level.

After the demographic questions, validated questionnaires were added to measure quality of life (8), delayed/cancelled medical care and productivity loss (9). In addition, several questions related to stress and concerns were also included, such as worries about the current and future financial situation, and concerns related to the national economy. Also, respondents were asked whether they had concerns about availability of medication, access to healthcare, or worries concerning individuals’ pre-existing conditions.

Quality of life was assessed by 2 instruments. First, a visual analogue scale was used to evaluate the overall health status of the participants on a scale from 0 to 100. The EuroQol 5 Dimensions questionnaire (10) measures health related quality of life on 5 dimensions (EQ-5D-5L) that map each dimension on a scale from below zero to 1. On that scale zero represents death, 1 represents perfect health and scores below zero reflect health states considered worse than death (8). The health state values are suitable for use in health economic evaluations(11). For the calculations, the Tobit with constraints model 3 was used with a constant of 0.953 (11). Respondents evaluated their health on the day of the questionnaire and before the lock-down. For both quality of life questionnaires respondents were asked to rate their health on the day of the questionnaire as well as the period before the lockdown.

Respondents answered questions related to medical resource use, cancelled or delayed health care appointments due to COVID-19 or COVID-19 related restrictions as assessed by the Medical Consumption Questionnaire (iMCQ) (12). This instrument was designed to measure medical resource use and includes questions related to the number of visits to various healthcare providers. Questions were adapted to be able to capture missed appointments to healthcare providers, including outpatient or primary care and inpatient or specialist care.

Productivity losses related to COVID-19 and COVID-19 related restrictions were recorded by the iMTA Productivity Cost Questionnaire (iPCQ) (9). The questions were slightly adapted to reflect the COVID-19 lockdown situation and included questions about employment status, absenteeism and presenteeism.

### Statistical analyses

The data were analyzed in IBM SPSS Statistics and summarized in proportions and means with 95% confidence intervals, and means with standard deviations when appropriate. The analyses were performed separately for Belgium and the Netherlands. In addition, subgroups of respondents were created based on age 18-35 years, 35-66 years (for Belgium) and 35-67 (for the Netherlands), and above pension age ≥66/≥67 years, respectively. We also categorized both countries by education level (low, middle, high), whereby low includes primary school, lower vocational education, preparatory secondary vocational education, middle includes higher general secondary education, secondary vocational education and high includes higher professional education, and university). Further subgroups were created by COVID-19 infection status including suspected of COVID-10 infection vs or not-infected); and by parent status (people with and without children below 12 years of age).

We calculated productivity costs related to lost paid work due to COVID-19 by multiplying the number of hours lost with the average age-related hourly income in the Netherlands (13) or Belgium (14). All costs are presented as weekly costs.

## Results

### Study population

A total of 2099 respondents completed the questionnaire in Belgium and 2058 in the Netherlands. The mean age was 46.4 year for Belgium and 47.6 year for the Netherlands (**Table 1**). Half of the respondents were women in both countries. In Belgium, 1.4% of the respondents tested positive by a confirmed COVID-19 test while this was 4.7% for the Netherlands. In both countries, similar proportions suspected that they might have been infected, without a confirmed test result (27% in Belgium and 22% in the Netherlands.

**Table 1:** Population characteristics by country

|  | Belgium (n=2099) | the Netherlands (n=2058) |
| --- | --- | --- |
| Age, year (SD) | 46.4 (16.3) | 47.7 (16.2) |
| Female (%) | 48.9 | 50.1 |
| Education level (%) |  |  |
| Low | 29.0 | 25.6 |
| Middle | 50.7 | 28.6 |
| High | 20.3 | 45.9 |
| Children, yes (%) | 57.5 | 60.7 |
| < 18 years | 23.8 | 29.2 |
| < 12 years | 15.5 | 22.1 |
| < 6 years | 9.1 | 13.5 |
| Tested for COVID-19 infection (%) | 6.3 | 8.8 |
| Tested positive (%) | 1.4 | 4.7 |
| Suspected COVID-19 infection |  |  |
| Yes/maybe (%) | 27.2 | 21.9 |

Values are proportions or mean and standard deviation; SD: Standard deviation

**Table 2:**
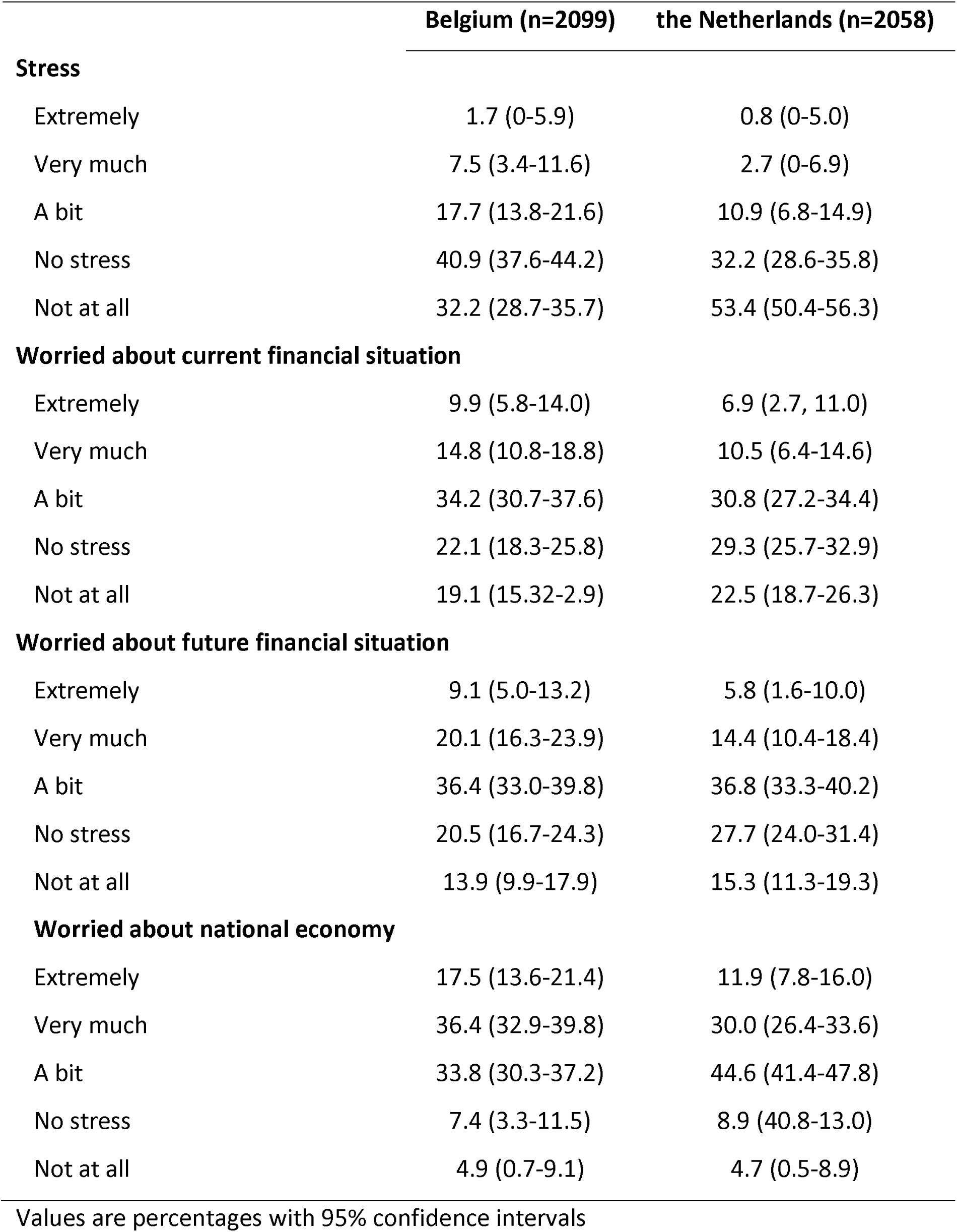
Stress and financial concerns during the first 8 weeks of the national COVID-19 measures

### Stress and concerns, quality of life

The results of the questions related to stress and financial concerns are presented in Error! Reference source not found.. A minority in both countries felt stressed (27% in Belgium and 14% in the Netherlands), but the majority reported concern about their personal current and future financial situation (59% and (48% respectively), and even more about the national economies (88% and 86%). Respondents reported good health, whether expressed on a visual analogue scale or by EQ-5D utilities (**Table 3**). Large proportions were worried about the national COVID-19 measures and about access to healthcare (**Table 4**).

**Table 3:** Perceived health and quality of life before and during the national COVID-19 measures

|  | **Belgium** | **the Netherlands** |
| --- | --- | --- |
|  | **(n=2099)** | **(n=2058)** |
| Perceived health during COVID-19 | 72.9 (71.0-74.0) | 73.1 (71.8-75.0) |
| Perceived health before COVID-19 | 74.5 (72.6-76.3) | 74.8 (72.9-76.7) |
| EQ-5D during COVID-19 measures | 0.79 (0.77-0.81) | 0.84 (0.82-0.86) |
| EQ-5D before COVID-19 measures | 0.82 (0.80-0.84) | 0.85 (0.83-87) |

Values are mean and 95% confidence intervals

**Table 4:**
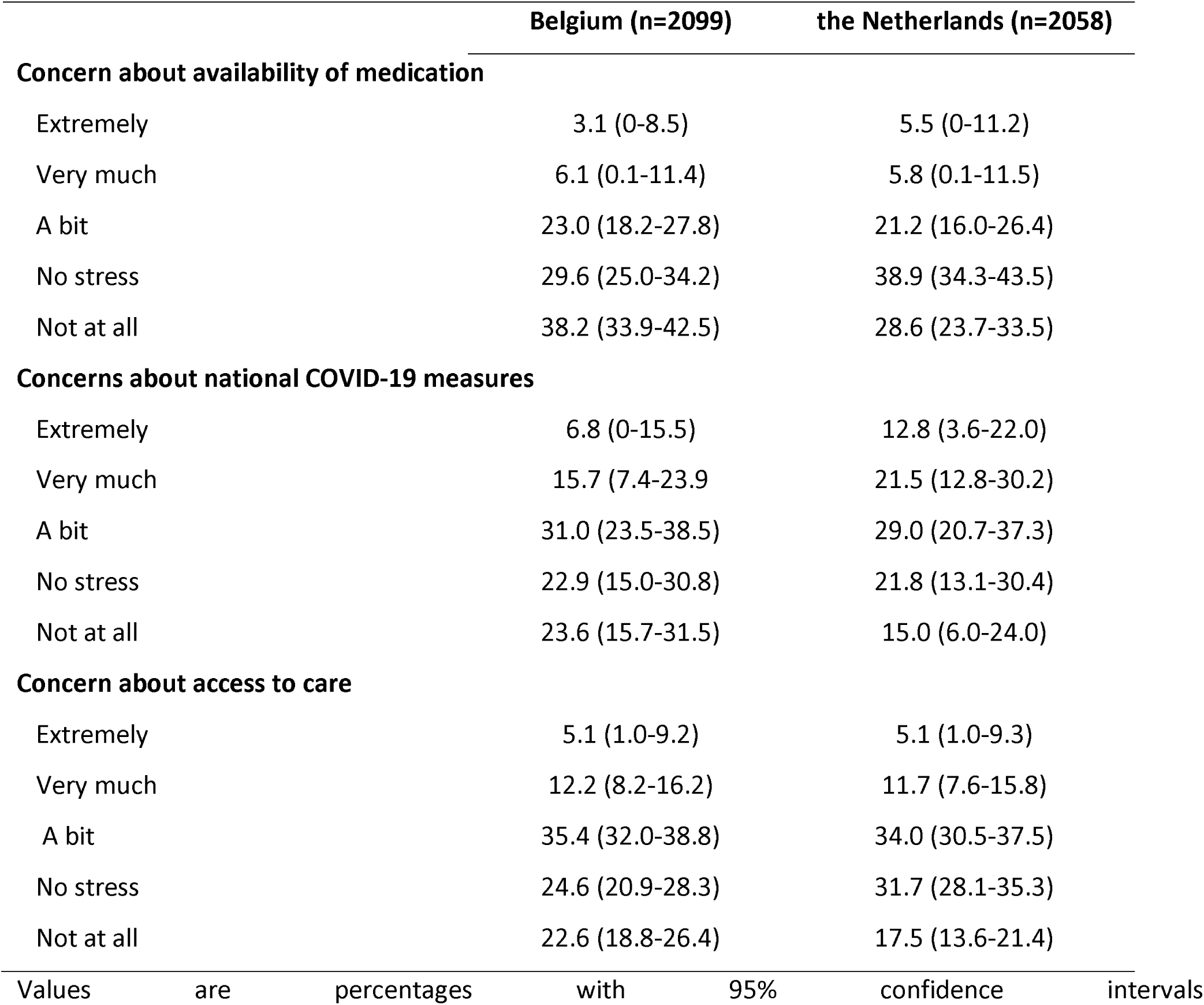
Concerns about care during the first 8 weeks of the national COVID-19 national measures

### Access to healthcare and cancelled/delayed healthcare

More than a quarter of respondents in both countries had experienced cancellations or postponements of appointments with healthcare providers (**Table 5**) while a higher percentage of respondents avoided a GP appointment. Most people agreed that they received the care (**Table 6**). Belgian respondents more often experienced medication supply issues (10.1%) vs the Dutch respondents (15.2%). The youngest age group (18-35 year) reported the highest medication supply issues (13.7%) vs (38.3%).

**Table 5:** Cancelled or postponed health-care by age groups

| Belgium |  |  |  |  | the Netherlands |  |  |  |
| --- | --- | --- | --- | --- | --- | --- | --- | --- |
|  | Total | 18- 35 year | 35-65 year | ≥ 66 year | Total | 18-35 year | 35-66 year | ≥ 67 year |
| N | 2099 | 641 | 1134 | 324 | 2058 | 560 | 1175 | 323 |
| **Experienced** |  |  |  |  |  |  |  |  |
| **cancelled/postponed** | 26.3 (24.4-28.2) | 28.7% (25.2-32.3) | 27.1 (24.5-29.7) | 19.1 (14.8-23.4) | 26.0 (24.1-27.9) | 31.8 (27.9-35.7) | 22.4 (20.0-24.8) | 19.1 (14.8-23.4) |
| **health-care** |  |  |  |  |  |  |  |  |
| N | 553 | 184 | 307 | 62 | 535 | 178 | 263 | 94 |
| **Avoided GP** |  |  |  |  |  |  |  |  |
| **appointment** | 51.9 (47.7-56.1) | 58.2% (51.1-65.3) | 51.8 (46.2-57.4) | 33.9 (22.1-45.7) | 45.0 (40.8-49.2) | 62.9 (55.8-70.0) | 40.3 (34.4-46.2) | 24.5 (14.1-34.9) |

Values are percentage with 95% confidence interval

**Table 6:** Issues with access to care and supply of medication by age groups

| Belgium |  |  |  |  | the Netherlands |  |  |  |
| --- | --- | --- | --- | --- | --- | --- | --- | --- |
|  | Total | 18- 35 year | 35-65 year | ≥ 66 year | Total | 18-35 year | 35-66 year | ≥ 67 year |
| N | 1265 | 291 | 715) | 259) | 1126 | 227 | 646 | 253 |
| Experienced medication supply issues |  |  |  |  |  |  |  |  |
|  | 10.1 (8.4-11.8) | 13.7 (9.7-17.7) | 10.6 (8.3-12.9) | 4.6 (2.0-7.2) | 15.2 (13.1-17.3) | 38.3 (32.0-44.6) | 10.8 (8.4-13.2) | 5.5 (2.7-8.3) |
| N | 2099 | 641 | 1134 | 324 | 2058 | 560 | 1175 | 323 |
| Received care as needed |  |  |  |  |  |  |  |  |
| Agree | 38.0 (34.6-41.4) | 35.9 (29.7-42.1) | 37.4 (32.8-42.0) | 44.8 (36.7-52.9) | 41.9 (38.6-45.1) | 44.5 (38.3-50.7) | 39.5 (35.1-43.9) | 46.1 (38.1-54.1) |
| Neutral | 21.2 (17.4-25.0) | 25.1 (18.4-31.8) | 19.9 (14.7-25.1) | 18.2 (8.4-28.0) | 19.4 (15.5-23.3) | 23.9 (16.7-31.1) | 18.8 (13.6-24.0) | 13.9 (3.8-24.0) |
| Disagree | 16.5 (12.6-20.4) | 16.7 (9.6-23.8) | 17.9 (12.6-23.2) | 10.7 (4.6-20.9) | 12.7 (8.7-16.7) | 11.8 (4.0-19.6) | 13.7 (8.4-19.0) | 11.5 (1.2-21.8) |
| Not applicable | 24.2 (20.5-27.9) | 22.3 (15.5-29.1) | 24.8 (19.7-29.8) | 26.2 (16.9-35.5) | 25.9 (22.2-29.6) | 19.8 (16.5-23.1) | 28.1 (25.5-30.7) | 28.5 (19.3-37.2) |

Values are percentage with 95% confidence interval

Paramedic care (physical therapist, dietician, social worker, psychologist) was cancelled more often than hospital care or 1^st^ line care (GP) in both Belgium and the Netherlands (**supplemental table 7**).

### Productivity loss

In total, 5% of the respondents in Belgium and 4% in the Netherlands reported that they lost their job due to COVID-19 **(Table 7)**. In Belgium higher productivity losses were reported for people who were employed compared to the Netherlands. More Belgian than Dutch respondents indicated that they were absent from work for at least one day (30% versus 19%). The mean value of lost production among respondents in paid profession per person per week including absenteeism and presenteeism was €161.39 for Belgium and €82.69 for the Netherlands.

**Table 7:** Productivity losses among respondents in paid profession Belgium

|  | Belgium | the Netherlands |
| --- | --- | --- |
| N | 1019 | 811 |
| Lost job due to COVID-19 (%) | 5.1 | 4.4 |
| N | 1080 | 1247 |
| In paid profession (%) | 52 | 61 |
| Worried about losing profession (%) |  |  |
| Somewhat to extremely | 39 | 40 |
| Not at all to slightly | 61 | 60 |
| Weekly work hours before COVID-19 | 34.4 (0.31.6-37.2) | 30.6 (28.0-33.2) |
| Weekly work hours during COVID-19 | 26.9 (24.3-29.5) | 26.6 (24.1-29.1) |
| Experienced absenteeism (%) |  |  |
| Some days | 16.5 | 9.6 |
| All days | 19.2 | 9.1 |
| Absenteeism (hours/week) | 1.6 (0.1-2.3) | 1.3 (0.1-1.9) |
| Cost absenteeism (person/week) | €133.86 | €63.58 |
| Experienced presenteeism (%) | 29.5 | 33.7 |
| Days experiencing presenteeism | 13.3 (11.3-15.3) | 9.6 (8.0-11.2) |
| Presenteeism (% of normal work per day) | 61% | 66% |
| Cost presenteeism (person/week) | €27.53 | €19.11 |

Values are mean and standard deviation or percentage with 95% confidence interval
\*Combined absenteeism for some days and all days

Subgroup analyses for age groups, parent status and COVID-19 infection are reported in supplemental tables 1-9.

## Discussion

This study described the stress and concerns and quality of life access to healthcare and cancelled/delayed healthcare productivity losses during the first 8 weeks of the coronavirus lockdown in Belgium and the Netherlands. Our study indicates that the respondents in Belgium and the Netherlands underwent considerable levels of stress and concerns and experienced cancelled/postponed care and productivity loss during the COVID-19 pandemic.

When comparing results between Belgium and the Netherlands, the respondents in Belgium were more worried than respondents in the Netherlands. Reported quality of life was lower in Belgium before and during the coronavirus lockdown. Additionally, both countries experienced similar levels of cancelled or postponed care as reported by the respondents. However, the respondents report different types of cancelled specialist care. For productivity loss, both countries reported similar levels of people losing their job due to COVID-19 and worries about losing your job. However, more people experienced productivity loss in Belgium compared to the Netherlands. This results in substantial higher costs related to presenteeism and absenteeism costs in Belgium than in the Netherlands. This could be due to temporarily installed paid unemployment leave regulations in Belgium.

The global scientific body of evidence is growing for stress, concerns, reduced quality of life, and productivity loss in relation to COVID-19. Several cross-sectional studies from varies countries assessed stress, quality of life, worries and various other questions related to the impact of COVID-19 with similar questionnaires (15, 16), particularly in China (17-19). A Chinese cross-sectional study reported that Chinese respondents have been greatly impacted in terms of non-work-related travel, work related travel, family’s daily routine, job and finance as well as reporting high levels of anxiety (17). On the other hand, another small Chinese study among highly educated participants reported mild stressful impact scores, however, 52% of that population felt horrified and apprehensive due to the pandemic (18). The majority of participants (58-78%) received increased support from friends and family members, increased shared feeling and caring with family members and others. A small study in Israel reported high levels of perceived stress and corona-related worries, but low levels of anxiety (15). Female sex, younger age, corona-related loneliness, and pre-existing chronic illness were all related to higher levels of psychological distress and lower levels of quality of life.

Another Chinese survey among university students, healthcare workers and business people indicated that during a 2-week period in February 2020, the emotional state, anxiety and behavior of participants in Hubei was lower compared to reference values in February 2019, particularly for sleep quality(19). These results from various countries underline the burden on our societies. Health education should be considered combined with psychological counseling for vulnerable individuals to cope with future outbreaks.

### Public health implications

The information gathered in this study gives a broad overview of the Belgian and Dutch society in the light of the COVID-19 pandemic and may be valuable for future health technology assessment studies in the field of COVID-19. It would be worthwhile to conduct a similar study at a later time in the pandemic, to investigate whether and how concerns, quality of life, access to care and productivity develop over time.

The current study highlights concerns, quality of life, and risk factors for psychological distress in light of the corona pandemic. More research is needed in order to fully understand the scope and correlates of societal difficulties during these challenging times.

### Strengths and limitations

Our study has some strengths and limitations. Strengths were the large representative sample that could be reached in a relatively short period of time, which was vital since the COVID-19 situation develops rapidly. We used validated questionnaires that strengthen the standardized data collection. Our population yielded a utility value of 0.85 for the entire population of the Netherlands, which is close to the reference value for the general Dutch population 0.87 (11). This implies that our sample is comparable to a general population sample.

A limitation of this study is that there was no ability to add detailed instructions to the questionnaire as only limited instructions could be provided at the beginning of the questionnaire. To obtain a broad view of society, the questionnaire was relatively long (mean duration 15 minutes) and the iPCQ and iMCQ can be considered quite complex. Some misunderstanding or mis categorization can therefore not be excluded. A second limitation is that the recall period in several sections of the questionnaire is relatively long (∼8 weeks). These responses maybe influenced by recall bias. The questions about the time before the lockdown relied on memory and were answered in a cross-sectional fashion. This might be influenced by external factors and might not represent normal life. Lastly, the sample was slightly underrepresented for the lowest educational class for Belgium.

## Conclusion

This study measured stress, quality of life, medical resource use and productivity losses in the general population in Belgium and the Netherlands during the first 8 weeks of the coronavirus lockdown. The results underline the burden on society in terms of stress, concerns, general healthcare and productivity.

## Data Availability

Data are available upon request.

## Acknowledgements

We thank all respondents for their valuable time and their participation. Furthermore, we would like to thank Lien Caveye and Kelly Mulder for their support.

## Data availability

Data are available upon request

